# Patient Screening for Self-Expanding Percutaneous Pulmonary Valves using Virtual Reality

**DOI:** 10.1101/2023.10.24.23297501

**Authors:** Jenny E. Zablah, Jeannie Than, Lorna P. Brownen, Salvador Rodriguez, Gareth J. Morgan

**Affiliations:** Heart Institute, Children’s Hospital Colorado. Department of Pediatrics University of Colorado Anschutz Medical Campus; Modern Human Anatomy School, University of Colorado Anschutz Medical Campus; Department of Radiology, University of Colorado Anschutz Medical Campus; Heart Institute, Children’s Hospital Colorado

**Author notes:** Corresponding Author: **Jenny Zablah MD**, <u>Email</u>, <u>Address</u>: 13123 E 16^th^ Avenue, Box 100, Aurora, Colorado, USA 80045.

## Abstract

**Background:** In recent years, self-expanding technology to treat pulmonary regurgitation in the native RVOT became FDA approved in the U.S. and is now routinely used. The current practice for selection of patients who are candidates for these devices includes screening for “anatomic fit”, performed by each of the manufacturing companies. Our study aims to validate the use of virtual reality as a tool for local physician-led screening of patients.

**Methods:** This retrospective study from Children’s Hospital Colorado included patients who underwent pulmonary valve replacement and had screening for a Harmony TPV or Alterra Pre Stent performed between September 2020 and January 2022. The data from the commercial companies dedicated analysis for self-expanding TPVR evaluation with perimeter analysis was collected. Virtual Reality simulation was performed blinded by two congenital interventional cardiologists using Elucis virtual reality software and an Oculus Quest 2 headset.

**Results:** Among the 27 evaluated cases, the use of a self-expandable valve was recommended by companies dedicated analysis in 23 cases (85.2%), by VR assessment in 26 cases (96.3) and finally implanted in 25 cases (92.6%). Regarding the level of agreement, both modalities (Manufacturer and VR) were good at screening *in* patients who received a self-expanding valve (100% vs. 96.1%). When it came to screening the patients *out*, VR presented good capacity to accurately classify non-suitable patients (50% vs. 100%).

**Conclusion:** Our institutional experience with virtual reality TPVR planning, accurately predicted clinical outcomes. This paves the way for routine use of VR in patient selection for self-expanding valve technologies.

## INTRODUCTION

Pulmonary valve dysfunction is common in patients with congenital heart disease (CHD), usually manifesting with right heart volume and/or pressure overload.^i^ Patients with pulmonary valve dysfunction may undergo multiple open-heart operations to avoid the consequences of right heart dysfunction. Transcatheter Pulmonary Valve Replacement (TPVR) has become an accepted non-surgical alternative for these patients, with a growing variety of prosthetic valves now available. In the last several years, self-expanding technology to facilitate native RVOT TPVR became FDA approved in the U.S. and is now routinely used. The two devices currently available are the Alterra Adaptive PreStent™ (Edwards Lifesciences, Irvine, CA) and the Harmony TPV (Medtronic, Minneapolis, MN). The Alterra Adaptive PreStent™ is a self-expanding, partially covered stent that was designed to internally reconfigure native RVOTs, making them suitable for implantation of a 29mm Edwards® SAPIEN S3 valve^ii^. The Harmony TPV is a porcine pericardial tissue valve mounted on a self-expanding nitinol frame. These are the only transcatheter devices designed specifically for the native RVOT that have FDA approval.

The current practice for selection of suitable patients for these devices include the clinical criteria for pulmonary valve replacement^iii^ and additional screening for *“anatomic fit”*. ECG gated cardiac computed tomography (CT) is performed, with the cardiac phase used for self-expanding TPVR evaluation depending on the valve type, as determined during the respective clinical trials (70% diastole for Harmony TPV and 30% systole for Alterra Adaptive PreStent™)^iv^. These images can be sent to one or both valve manufacturers, where a perimeter plot with a valve fit analysis is performed and sent back to the physician implanter with recommendations of potential suitability in each case.

Virtual Reality (VR) is a tool that has gained popularity for procedural planning.^v^ Using VR for procedural simulations requires specific software and equipment to allow accurate and consistent results. Once this is in place and a reliable clinical workflow established, simulations can be produced in minutes, allowing spatial conceptualization of complex anatomical structures, and giving the capability to view and interact with dynamic models in a 3D virtual space using patient-specific imaging datasets^vi^.

Our study aims to validate the use of virtual reality as a tool for physician led screening of patients with clinical criteria for TPVR. We planned to determine suitability for the available self-expanding technologies and compare this determination with both the manufacturer screening reports as well as the clinical outcomes of these patients.

## MATERIALS AND METHODS

This retrospective study was performed at Children’s Hospital Colorado. Patients who underwent percutaneous pulmonary valve replacement who had previously undergone screening for either a Harmony TPV or Alterra Pre Stent between September 2020 and January 2022 were included in the study.

### Imaging

A database of patients that had undergone screening with a retrospectively ECG gated cardiac computed tomography (CT) scan for Alterra and Harmony was collected. The CTs were exported onto Circle Cardiovascular Imaging software to compress and export the files in an anonymized DICOM format for processing and segmentation in a virtual reality environment.

### Modeling

Elucis (Realize Medical, Inc. Canada), an FDA approved virtual reality software, was used to create the 3D models, and perform the simulations. The DICOMs created from each patient’s CT scans were uploaded onto Elucis. A 3-D model was created by adjusting Hounsfield thresholds of the anatomy. Each model included the left and right pulmonary arteries, main pulmonary artery, RVOT, and right ventricle to ensure the presence of adequate anatomical structures to help identifying a potential landing zone for each valve. Images of the segmentation steps can be viewed in Figure 1.

**Figure 1.**
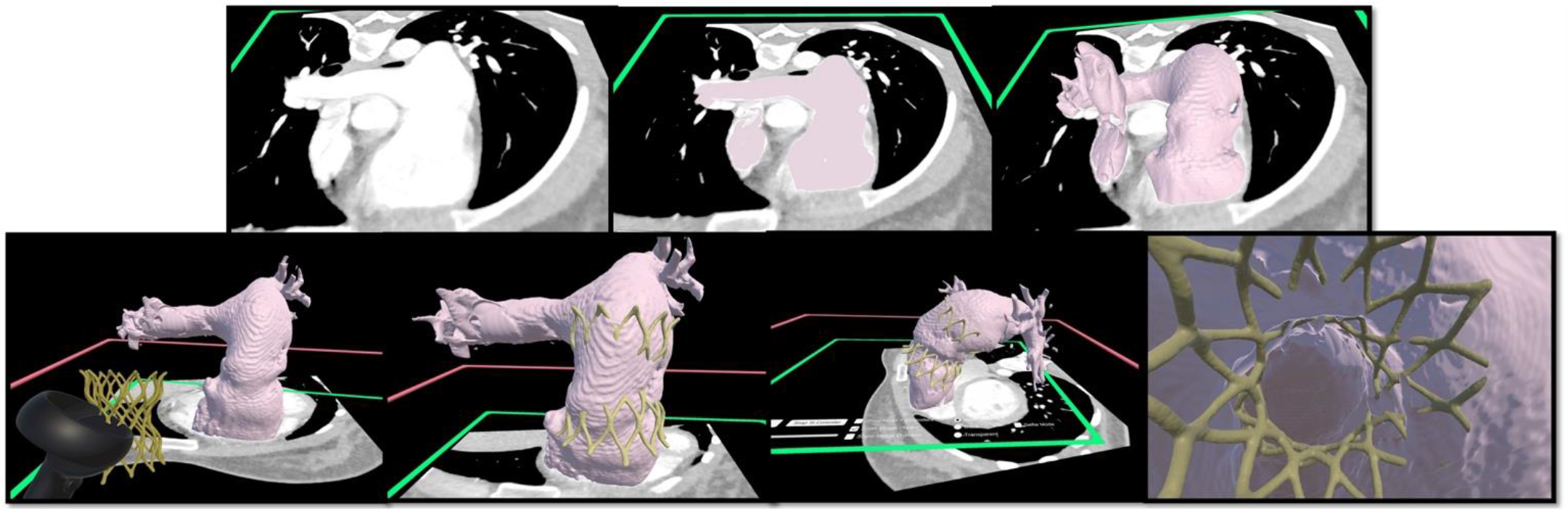
Steps of creating a 3D model of the RVOT using Elucis (Realize Medical Inc., Canada). A DICOM file of a dynamic CTA is loaded into the 3D environment of Elucis. The CT images can be viewed in 2D as in any DICOM viewer and then using thresholding, the area of interest is selected. A 3D model is then created in a 2D view (panel B) or in a 3D view (panel C). This 3D model is now dynamic and will show the MPA characteristics during the cardiac cycle. The models of the different valves evaluated are imported and placed in the area of interest to simulate the implantation position. The views include internal aspects of the stent and outflow tract to assess anatomical interaction between the two.

The models of the Alterra Adaptive PreStent and Harmony TPV utilized for simulation were created by performing a 3D rotational angiography of the valve frames, which were then segmented, and saved as STL files. The STL file was then imported to coexist in the virtual environment of the dynamic 3D cardiac reconstruction. Manipulation of both these structures in the same 3D space was then used to rehearse multiple scenarios, planning the potential options for the actual clinical case (Figure 2).

**Figure 2.**
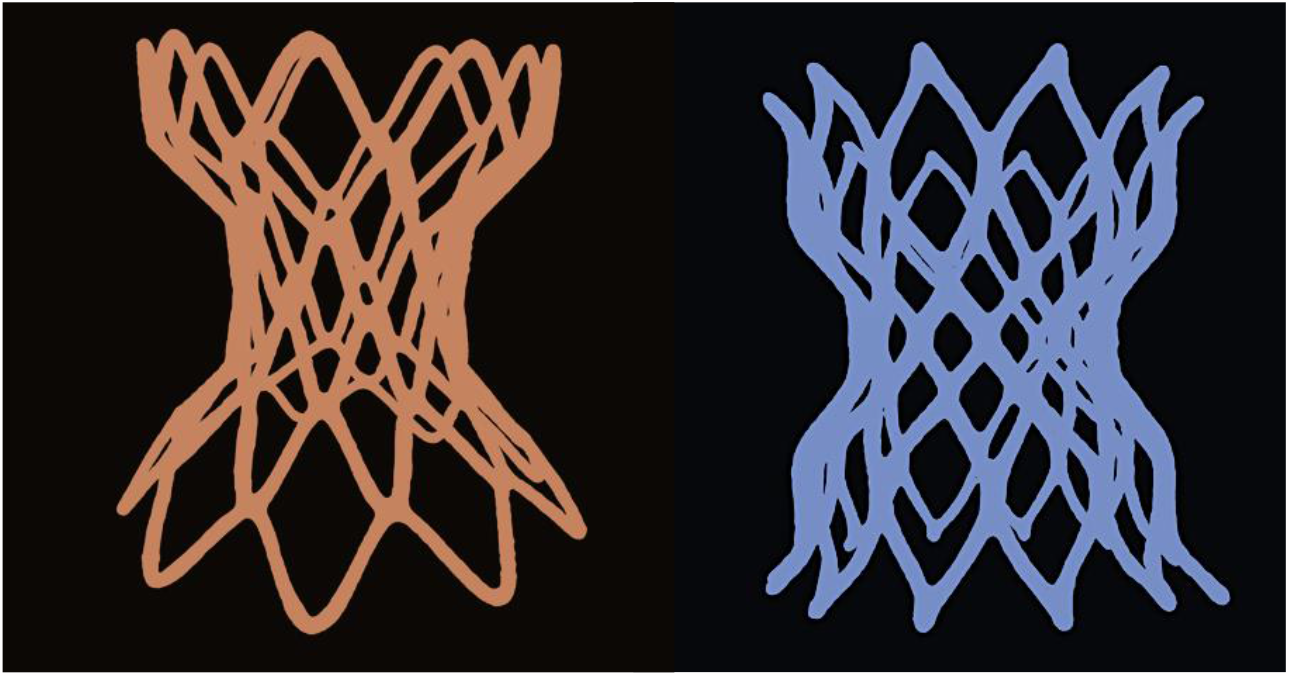
3D models created by our team utilizing 3D rotational angiography of the devices at scale. The resulting STL files were used to simulate valve implantation within Elucis.

### Data Collection

Retrospective chart review was performed to collect demographic data of the patients. The data from each manufacturers dedicated analysis for self-expanding TPVR evaluation with perimeter plots to determine anatomic fit was collected and reviewed.

Simulation was performed by two congenital interventional cardiologists from the cardiology catheterization laboratory at Children’s Hospital Colorado using the Elucis virtual reality software (Realize Medical) and Oculus Quest 2 headset. Unidentified dynamic 3D models (through systole and diastole) were shown in the virtual environment and simulated live implantation of the imported Harmony TPV and Alterra Adaptive PreStent valves were performed to predict the valve best fit for the patient. Each implanter performed the simulation independently to ensure their predictions were not influenced by outside factors such as collaboration from other physicians. The interventional cardiologists were blinded to the actual clinical outcome of the cases. 3D models of the Harmony TPV 25mm, Harmony TPV 22mm, Alterra Adaptive PreStent and 29mm Edwards Sapien 3 valve were used in every case for the live simulation.

### Analysis

Statistical analyses were performed using SPSS 28 (IBM software group, Chicago, Illinois). Categorical data were summarized by frequency and percentage. Continuous data were shown as median and range. Cohen’s kappa index was used to determine the level of agreement between companies’ recommendation, VR assessment, and the final clinical outcome (whether the valve was placed or not). A *p*-value of 0.05 was established as significant.

## RESULTS

Patients in this cohort were predominantly male (59.3%) with a median age of 28 years (range 12-54 years), and Tetralogy of Fallot was the most common diagnosis (59.3%). The rest of demographic characteristics are summarized in Table 1.

**Table 1.** Demographic characteristic.

| Table 1. Demographic characteristic |  |
| --- | --- |
| **Sex** |  |
| Male | 16 (59.3%) |
| Female | 11 (40.7%) |
| Age, years | 28 (12 – 54) |
| Weight, Kg | 70.3 (41 – 112) |
| Height, cm | 168 (122 – 190) |
| BSA, kg/m<sup>2</sup> | 1.8 (1.37 – 2.33) |
| BMI, kg/m<sup>2</sup> | 24.9 (16.8-49.4) |
| **Cardiac diagnosis** |  |
| Tetralogy of Fallot | 16 (59.3%) |
| Congenital Pulmonary Valve Stenosis | 11 (40.7%) |
| Values are shown as medians and ranges (min - max) or numbers as a percentage of the cohort (%). |  |
| BSA = Body surface area. |  |
| BMI = Body Mass Index |  |

Among the 27 evaluated cases, the use of self-expandable valve was recommended by the manufacturers dedicated analysis in 23 cases (85.2%), by VR assessment in 26 cases (96.3) and finally implanted in 25 cases (92.6%). The recommendations for each case are summarized in Table 2.

**Table 2.** Companies and VR recommendation in relation to Clinical outcome.

|  |  |  |  |
| --- | --- | --- | --- |
| Case Number | Did companies screen this patient *in*? | VR assessment screened the patient *in*? | Was a self-expanding valve placed in this patient? |
| VR-1 | Yes | Yes | Yes |
| VR-2 | Yes | Yes | Yes |
| VR-3 | No | Yes | Yes |
| VR-4 | Yes | Yes | Yes |
| VR-5 | Yes | Yes | Yes |
| VR-6 | Yes | Yes | Yes |
| VR-7 | Yes | Yes | Yes |
| VR-8 | Yes | Yes | Yes |
| VR-9 | Yes | Yes | Yes |
| VR-10 | No | Yes | Yes |
| VR-11 | Yes | Yes | Yes |
| VR-12 | Yes | Yes | Yes |
| VR-13 | Yes | Yes | Yes |
| VR-14 | Yes | Yes | Yes |
| VR-15 | Yes | Yes | Yes |
| VR-16 | Yes | Yes | Yes |
| VR-17 | Yes | Yes | Yes |
| VR-18 | Yes | Yes | Yes |
| VR-19 | Yes | Yes | Yes |
| VR-20 | Yes | Yes | Yes |
| VR-21 | Yes | Yes | Yes |
| VR-22 | No | No | No |
| VR-23 | Yes | Yes | Yes |
| VR-24 | No | Borderline (H) | No |
| VR-25 | Yes | Yes | Yes |
| VR-26 | Yes | Yes | Yes |
| VR-27 | Yes | Yes | Yes |
| H-Harmony TPV 25mm; A-Alterra Adaptive PreStent; S3-Edwards Sapien S3, Sx: Surgical |  |  |  |

When recommendations were compared to the final clinical outcome, there was a substantial level of agreement between the screening tools used by companies and the final clinical outcome (Cohen’s Kappa 0.630, *p* <0.001); the recommendation generate using VR presented a slightly higher level of agreement to the final clinical outcome (Cohen’s Kappa 0.649, *p* <0.001).

Both modalities (companies and VR) were good at screening *in* patients who eventually received a self-expandable valve (100% vs. 96.1%); but the accuracy of screening patients *out*, was higher with the VR screening when compared with manufacturer data (50% vs. 100%). These percentages are summarized in Table 3, and discrepancies are summarized in Table 4.

**Table 3.** Level of Agreement Between Screening Approval and successful Clinical Valve implantation.

|  |  |  |  |  |  |
| --- | --- | --- | --- | --- | --- |
| Groups | Self-expandable valve | Was placed<br>n = 25 | Was **not** placed<br>n = 2 | Cohen’s Kappa | *p* value |
| Companies’ systems | Was recommended | 23 (100%) | 0 (0%) | 0.630 | <0.001 |
|  | Was not recommended | 2 (50%) | 2 (50%) |  |  |
| VR testing | Was recommended | 25 (96.2%) | 1 (3.8%) | 0.649 | <0.001 |
|  | Was not recommended | 0 (0%) | 1 (100%) |  |  |

**Table 4.** Analysis of Case Discrepancies.

| Table 4. Analysis of Case Discrepancies |  |  |  |  |  |  |
| --- | --- | --- | --- | --- | --- | --- |
| *Cases* | Clinical Outcome | Company | VR |  |  | Comments |
|  |  |  | Overall | Ev. 1 | Ev. 2 |  |
| *Case 3* | Yes | No | Yes | Yes | Yes | Borderline large MPA size |
| *Case 10* | Yes | No | Yes | Yes | Yes | Borderline large MPA size |
| *Case 24* | No | No | No | No | Yes | Patient was 20kg and short MPA. Decision was made to go to surgery. |

Regarding the diagnostic performance of both modalities (companies and VR), both tools presented a high level of sensitivity (92% vs. 100%) and positive predictive value (100% vs. 96.2%) but had less consistent results in relation to specificity (100% vs. 50%) and negative predictive value (50% vs. 100%). Overall diagnostic accuracy was high using both screening methods (92.5% vs. 96.3%).

## DISCUSSION

Self-expanding technology for TPVR has allowed us to capture patients with dysfunctional pulmonary valves that previously would be referred for surgical repair. Identifying the appropriate patients with suitable outflow tract and pulmonary artery anatomy for TPVR is critical for procedural success and appropriate patient counseling and education^vii^.

Our study demonstrated that virtual reality can be a useful tool for manufacturer agnostic screening of these patients. It facilitates the understanding each individual patient’s anatomy and procedure planning based on case simulation. In the study, all patients that were accepted for TPVR by the manufacturer analysis were also accepted by VR^viii^. Interestingly, there were two patients that were screened *out* by the companies that were screened *in* by VR. These patients underwent successful TPVR with self-expanding platforms. This was an interesting finding that is likely multifactorial. The manufacturer led screenings were performed early in their experience with self-expanding valves and as time and experience has increased, physicians and industry screening protocols have started to challenge complex anatomy more. The physicians in our study are both very experienced TPVR operators, which may make them more likely to push the envelope when it comes to patient inclusion. However, surely the learning environment which VR engenders during each patient simulation provides an iterative progressive platform through which the physicians understanding of the interaction between anatomy and implant blossoms. This has an unquantified but definite impact on the physician’s ability to see a path to procedural success through complex anatomy.

Successful simulation of cases like these, requires appropriate software but also the need for the physician to feel comfortable manipulating models, understanding the interactions of the valve frames with the anatomy and interpreting the dynamic images accordingly. Having imaging background or interest help to develop skills to use this technology, but also using the technology routinely, allows more comfort and confidence with it. Having a team member with specialized interest in creating and interpreting these models is a good approach to allow consistent and safe outcomes.

In our center, initial screening is performed in most cases using VR to identify patients that may need the ECG gated CTA for official manufacturer-based screening. Routine simulation of these cases in our center, allows implanters a different level of comfort and knowledge of the anatomy which reflects in the level or preparedness for each case and hopefully, overall outcomes.

We demonstrate that our institutional experience with virtual reality TPVR planning matches the clinical outcomes and is at least on a power with manufacturer-based screening. This preliminary study should open the door for cautious adoption of VR as a routine tool for patient selection for self-expanding valve platform.

## Data Availability

All data is available per request

## FUNDING

Project K: Legacy Grant from the World’s Children’s Initiative (WCI). In memory of Dr Kanishka Ratnayaka.

## Notes

### Competing Interest Statement

Dr Gareth Morgan and Dr Jenny Zablah are consultants for Edwards Lifescience and for Medtronic.

### Author Declarations

University of Colorado COMIRB

## References

i Cools B, Brown SC, Heying R, Jansen K, Boshoff DE, Budts W, Gewillig M. Percutaneous pulmonary valve implantation for free pulmonary regurgitation following conduit-free surgery of the right ventricular outflow tract. Int J Cardiol. 2015; 186:129-35. doi: 10.1016/j.ijcard.2015.03.108. Epub 2015 Mar 10. PMID: 25818755.

ii Zablah, J. E., Morgan, G. J. (2020a). Current Treatment Options for Catheter-Based Pulmonary Valve Replacement in Children. Current Treatment Options in Pediatrics, 6(4), 274–282. 10.1007/s40746-020-00209-0.

iii Ran L, Wang W, Secchi F, Xiang Y, Shi W, Huang W. Percutaneous pulmonary valve implantation in patients with right ventricular outflow tract dysfunction: a systematic review and meta-analysis. Ther Adv Chronic Dis. 2019 Jun 14;10:2040622319857635. doi: 10.1177/2040622319857635. PMID: 31236202; PMCID: PMC6572891.

iv Neil D. Patel, Daniel S. Levi, John P. Cheatham, Shakeel A. Qureshi, Shabana Shahanavaz, Evan M. Zahn, Transcatheter Pulmonary Valve Replacement: A Review of Current Valve Technologies, Journal of the Society for Cardiovascular Angiography & Interventions, Volume 1, Issue 6, 2022, 100452, ISSN 2772-9303, 10.1016/j.jscai.2022.100452.

v Ghosh, R.M., Jolley, M.A., Mascio, C.E., Chen, J.M., Fuller, S., Rome, J.J., Silvestro, E. and Whitehead, K.K., 2022. Clinical 3D modeling to guide pediatric cardiothoracic surgery and intervention using 3D printed anatomic models, computer aided design and virtual reality. 3D Printing in Medicine, 8(1), p.11.

vi Zablah, J. E., Morgan, G. J. (2020b). Innovations in Congenital Interventional Cardiology. The Pediatric clinics of North America, 67(5), 973–993. 10.1016/j.pcl.2020.06.012.

vii Kang, Sok, Leng, Benson, Lee. (2018). Percutaneous pulmonary valves for the large outflow tract [Figure]. Recent advances in cardiac catheterization for congenital heart disease [version 1; peer review: 2 approved]. F1000 Research, 7, 370.

viii Redondo, A., Baldrón, C., Aguiar, J.M., Juanatey, J.R.G., San Román, A. and Amat-Santosa, I.J., Use of extended realities in interventional cardiology: mixed reality for TAVI procedure.

